# Recreational and occupational physical activity and risk of adverse events in truncating *MYBPC3* founder variant carriers

**DOI:** 10.1101/2024.01.10.24301022

**Authors:** Fahima Hassanzada, Mark Jansen, Freyja H.M. van Lint, Laurens P. Bosman, Amand F. Schmidt, Dennis Dooijes, Danny van de Sande, Bristi Miah, Saskia N. van der Crabben, Arthur A.M. Wilde, Ronald H. Lekanne Deprez, Rudolf A. de Boer, Imke Christiaans, Jan D.H. Jongbloed, Harald T. Jorstad, Folkert W. Asselbergs, J. Peter van Tintelen, Annette F. Baas, Anneline S.J.M. te Riele

## Abstract

**Background:** *MYBPC3* founder variants cause hypertrophic cardiomyopathy (HCM) leading to heart failure (HF) and malignant ventricular arrhythmias (MVA). Exercise is typically regarded a risk factor for disease expression, although evidence is conflicting. Stratifying by type of exercise may discriminate low-from high-risk activities in these patients.

**Objective(s):** Evaluate effects of exercise, stratified by high-static and high-dynamic component, on risk of major cardiomyopathy-related events (MCE) and cardiomyopathy penetrance among *MYBPC3* founder variant carriers.

**Methods:** We interviewed 188 carriers (57% male, 43.4±14.8 years) on exercise participation since age ten. Exercise was quantified as metabolic equivalent task (MET)-hours/week before presentation. MCE was defined as a composite of MVA (sustained ventricular tachycardia/fibrillation), HF (HF hospitalizations or transplantation), and septal reduction therapy. Static and dynamic exercise were defined per Bethesda classification. Associations of exercise with MCE and cardiomyopathy penetrance were adjusted for sex and assessed using Cox regression.

**Results:** Overall, 40 (21%) subjects experienced MCE and 139 (74%) were diagnosed with cardiomyopathy. No association was found for overall physical activity and high-static activity with MCE (p=0.900 overall; p=0.274 high-static) or cardiomyopathy penetrance (p=0.787 overall; p=0.774 high-static). In contrast, high-dynamic activity was associated with MVA (dichotomized at 75^th^ percentile: adjusted hazard ratio 2.70, 95% confidence interval 1.01-7.24, p=0.049).

**Conclusions:** Overall exercise participation does not generally increase the risk of adverse events among *MYBPC3* founder variant carriers. Nonetheless, an increased risk of MVA was observed among those engaged in the highest quartile of high-dynamic-sports, suggesting that high-level high-intensity exercise activities should be entertained with caution.

## Introduction

Hypertrophic cardiomyopathy (HCM) is a relatively common inherited heart disease with an estimated prevalence of 1:500.^1^ It is characterized by left ventricular hypertrophy which is not explained by abnormal loading conditions.^2^ Clinical presentation is heterogeneous with many patients remaining mildly symptomatic or asymptomatic, while others present with left ventricular (LV) outflow tract obstruction requiring septal reduction therapy (SRT), malignant ventricular arrhythmias (MVA) or debilitating heart failure (HF). This heterogeneity suggests an influential role of environmental factors.^3^

HCM is predominantly caused by variants in genes encoding sarcomeric proteins. In approximately 50% of patients, a likely pathogenic or pathogenic variant is found.^4,5^ Variants are most frequently found in the *MYBPC3* gene, encoding cardiac myosin-binding protein C, an important regulator of cardiac contractility.^6^ In the Netherlands, approximately 20% of HCM patients carry a truncating *MYBPC3* founder variant.^7^ These variants lead to truncation of *MYBPC3* mRNA and subsequent haploinsufficiency of cardiac myosin-binding protein C, which has been shown to impact prognosis similar to other truncating *MYBPC3* variants.^8,9^

As HCM has frequently been reported in young athletes suffering sudden cardiac death, guidelines have traditionally discouraged competitive exercise in HCM patients.^10–12^ Exercise is hypothesized to contribute to sudden cardiac death in HCM by several mechanisms, including increasing LV wall thickness and outflow tract obstruction, inducing metabolic disturbances or ischemia, as well as pro-arrhythmic effects by adrenergic stimulation and/or fibrosis formation.^12,13^ However, several studies investigating effects of exercise in HCM have not reported negative effects, with a single study even showing a protective effect on all-cause and cardiovascular mortality.^14–16^ Of note, none of these studies assessed specific types of exercise, such as a distinction between static and dynamic properties.^15^ High-static exercise, for example, may theoretically be more likely to contribute to hypertrophic remodeling, compared with high-dynamic exercise.^17,18^

Therefore, we evaluated the effect of recreational and occupational physical activity on risk of major cardiomyopathy-related events (MCE) and cardiomyopathy penetrance in carriers of truncating *MYBPC3* founder variants, stratified by high-static and high-dynamic properties.

## Methods

### Patient population

This study is embedded in the BIO For CARe (Identification of BIOmarkers of HCM development and progression in Dutch *MYBPC3* Founder variant CARriers) and UNRAVEL (www.unravelrdp.nl) registries.^19,20^ Carriers of the c.2373dupG, c.2827C>T, c.2864_2865delCT and c.3776delA *MYBPC3* founder variants were recruited from three Dutch university medical centers (Utrecht, Groningen and Amsterdam). The Institutional Review Board, Medical Ethics Committee of UMC Utrecht gave ethical approval for this work, with local approval at the Amsterdam UMC and UMC Groningen. All individuals provided written informed consent.

### Clinical data and outcomes

Clinical data were obtained from electronic health records and collected using pre-specified electronic case report forms, as previously described.^19^ Collected data included sex, date of birth, date and type of clinical presentation, hospitalization records, and results of (non-) invasive studies (12-lead electrocardiogram [ECG], 2-dimensional transthoracic echocardiography, and cardiac magnetic resonance imaging). Clinical presentation was defined as the first medical visit for cardiac screening and/or symptoms related to *MYBPC3* in one of the enrolling centers.

The primary outcome was a composite of MCE, constituting of MVA (defined as sustained ventricular tachycardia, defined as ≥30 seconds at ≥100 beats per minute and resulting in hemodynamic instability or requiring termination, aborted cardiac arrest or sudden cardiac death), HF events (defined as HF hospitalization, ventricular assist device implantation, heart transplantation or HF-related mortality), and SRT (defined as septal myectomy or alcohol septal ablation).

Secondary outcomes were MVA, HF events, and SRT, analyzed as separate outcomes. In addition, we evaluated the development of an overt cardiomyopathy diagnosis as secondary outcome measure. For the latter, HCM was defined as a maximum left ventricular wall thickness of ≥13 mm not explained by abnormal loading conditions;^11^ dilated cardiomyopathy (DCM) as dilation of left ventricle with a left ventricular ejection fraction (LVEF) <45%;^21^ Non-compaction cardiomyopathy (NCCM) as left ventricular non-compaction fulfilling the Jenni or Petersen criteria and a LVEF <50%,^22,23^ all not explained by abnormal loading conditions or coronary artery disease. Individuals with LVEF <50% but without other criteria for a specific cardiomyopathy diagnosis were classified as cardiomyopathy not otherwise specified (CMP-NOS).

### Physical activity interviews

Structured telephone interviews were conducted by three research assistants and one physician. Details of the exercise interview have been described previously.^24,25^ In short, physical activity was documented since age 10 years onwards using a pre-defined data collection sheet for all recreational and occupational physical activity. For each entry, the intensity (light, moderate or vigorous) and duration of physical activity were registered. Subsequently, the corresponding metabolic equivalents (METs) were assigned using the Ainsworth et al. compendium of physical activities.^26^

We assessed the impact of physical activity from age 10 years until presentation by stratifying in: (i) overall physical activity; and (ii) physical activity during high-static and high-dynamic sports. Overall physical activity was quantified as the average MET-hours/week spent on any physical activity and analyzed both in quartiles (based on the data) and based on increments of 7.5 MET-hours/week (which is the American Heart Association’s recommended minimum physical activity) to compare physical activity to standardized recommendations. High-static and high-dynamic sports were classified according to the 36^th^ Bethesda Conference Classification of Sports (Supplemental Figure 1) and quantified as MET-hours/week spent on this specific activity.^17,27^ In this study, sport activities in categories IIIA, IIIB and IIIC were classified as high-static; and sport activities in categories IC, IIC and IIIC as high-dynamic. Of note, class IIIC overlaps in both high-static and high-dynamic categories. As high-static and high-dynamic activity categories were analyzed separately, the use of class IIIC in both categories was statistically permitted.

### Statistical analysis

Analyses were performed in R software (version 4.2.2). Continuous variables were expressed as mean ± standard deviation or median [interquartile ranges (IQR)] as appropriate and compared using independent samples t-tests or Mann-Whitney U tests, respectively.

Categorical variables were presented as frequencies (percentage) and compared using the chi-square or Fisher’s exact test. Physical activity before and after presentation were compared using the Wilcoxon signed rank test.

Cumulative freedom from the composite outcome was determined by the Kaplan-Meier method and Cox-proportional hazard model. Follow-up was considered to start at first clinical presentation and end at the date of reaching the endpoint or censoring (i.e. death from any other cause or most recent follow-up at which the endpoint could be ascertained). Schoenfeld residuals test was used to check the proportional hazard assumption. The linearity assumption in the Cox model was assessed using Martingale residuals. In the absence of established cut-off points for sport class-specific activities, medians and 75^th^ percentile were used to dichotomize activities in case of significant non-linearity.^28^ Multivariable Cox regression was performed to correct for potential confounding from sex. Sensitivity analyses were performed by restricting the analyses to patients diagnosed with a cardiomyopathy diagnosis at first presentation. P-values <0.05 were considered statistically significant throughout.

## Results

### Study population

We included 188 carriers of truncating *MYBPC3* founder variants. Their clinical characteristics are provided in Table 1. Overall, 108 (57%) were male, with a mean age 43.4±14.8 years at first presentation. Almost half (n=80, 43%) of study subjects were probands. Maximum wall thickness was 16±6 mm with a mean LV ejection fraction of 59% ±10 and a LV end-diastolic diameter of 46±6 mm. At first clinical evaluation, 104 (55%) had a diagnosis of cardiomyopathy, most commonly HCM (n=98, 52% of overall population) (Supplemental Table 1).

**Table 1.** Baseline characteristics stratified by quartiles of overall physical activity*.

|  | **Overall**<br>**(n=188)** | **Quartile 1**<br>**(n=47)** | **Quartile 2**<br>**(n=47)** | **Quartile 3**<br>**(n=47)** | **Quartile 4**<br>**(n=47)** | **p-value** |
| --- | --- | --- | --- | --- | --- | --- |
| **Male sex** | 108 (57) | 21 (45) | 23 (49) | 26 (55) | 38 (81) | 0.002 |
| **Age at presentation (years)** | 43.4±14.8 | 42.6±13.2 | 42.1±16.7 | 44.2±15.0 | 44.5±14.4 | 0.830 |
| **Proband** | 80 (43) | 19 (40) | 16 (34) | 20 (43) | 25 (53) | 0.301 |
| **Maximum wall thickness (mm)** | 16±6 | 15±7 | 16±7 | 16±6 | 16±6 | 0.968 |
| **LV ejection fraction (%)** | 59±10 | 60±7 | 59±9 | 60±8 | 57±13 | 0.552 |
| **LV end-diastolic diameter (mm)** | 46±6 | 44±5 | 46±7 | 45±6 | 48±7 | 0.157 |
| **Left atrial diameter (mm)** | 40±7 | 39±6 | 40±7 | 40±10 | 40±6 | 0.962 |
| **Body surface area (m<sup>2</sup>)** | 2.0 [1.8-2.1] | 1.9 [1.8-2.1] | 1.8 [1.8-2.1] | 2.0 [1.7-2.1] | 2.0 [1.9-2.1] | 0.786 |

\*Subject characteristics, stratified by average MET-hours/week of recreational and occupational physical activity quartiles before first clinical presentation; (Quartile 1: 0-25 MET-hours/week); (Quartile 2: 26-56 MET-hours/week), (Quartile 3: 57-110 MET-hours/week); (Quartile 4: 113- 351 MET-hours/week). Values are shown as mean ± standard deviation, median (interquartile range) or counts (percentage). Abbreviations as in text.

**Table 2.** Associations of physical activity with primary endpoint (major cardiomyopathy-related events)

|  | **aHR\*** | **95% CI** | **p-value** |
| --- | --- | --- | --- |
| Overall physical activity (per 7.5 MET-hours/week of AHA minimum recommended exercise level) | 1.00 | 1.00-1.00 | 0.900 |
| High dynamic (per 1 MET-hours/week) | 1.00 | 0.98-1.02 | 0.826 |
| High static (per 1 MET-hours/week) | 0.97 | 0.92-1.02 | 0.274 |

\*Corrected for sex. Abbreviations as in text.

**Table 3.** Associations of physical activity with secondary endpoints (MVA, HF events, SRT and cardiomyopathy penetrance)*

|  | **aHR<sup>†</sup>** | **95% CI** | **P-value** |
| --- | --- | --- | --- |
| **MVA** |  |  |  |
| Overall physical activity (per 7.5 MET-hours/week of AHA minimum recommended exercise level) | 1.00 | 1.00-1.00 | 0.422 |
| High-dynamic (dichotomized by median 10 MET-hours/week) | 2.29 | 0.80-6.55 | 0.122 |
| High-dynamic (dichotomized by 75<sup>th</sup> percentile 23 MET-hours/week) | 2.70 | 1.01-7.24 | 0.049 |
| High static (per 1 MET-hours/week) | 0.98 | 0.92-1.05 | 0.589 |
| **HF events** |  |  |  |
| Overall physical activity (per 7.5 MET-hours/week of AHA minimum recommended exercise level) | 1.00 | 1.00-1.00 | 0.869 |
| High-dynamic (per 1 MET-hours/week) | 1.01 | 0.99-1.04 | 0.338 |
| High-static (per 1 MET-hours/week) | 0.95 | 0.83-1.08 | 0.432 |
| **SRT** |  |  |  |
| Overall physical activity (per 7.5 MET-hours/week of AHA minimum recommended exercise level) | 1.00 | 1.00-1.00 | 0.810 |
| High-dynamic (per 1 MET-hours/week) | 0.97 | 0.93-1.01 | 0.169 |
| High-static (per 1 MET-hours/week) | 0.98 | 0.91-1.06 | 0.661 |
| **Cardiomyopathy penetrance** |  |  |  |
| Overall physical activity (per 7.5 MET-hours/week of AHA minimum recommended exercise level) | 1.00 | 1.00-1.00 | 0.787 |
| High-dynamic (per 1 MET-hours/week) | 0.99 | 0.97-1.01 | 0.259 |
| High-static (per 1 MET-hours/week) | 1.00 | 0.97-1.02 | 0.774 |

\*Associations were preferentially analyzed as continuous values and presented as such, or presented using clinically meaningful cutoffs (e.g. recommended minimum of overall activity). For high-dynamic exercise and the MVA outcome, however, the linearity assumption was violated and data is presented in a dichotomized fashion. † Corrected for sex. Abbreviations as in text.

### Physical activity history

Before first medical contact, study participants were physically active for a median of 56 [26-111] MET-hours/week, of which 15 [6-29] MET-hours were spent on recreational activity, and 34 [4-84] MET-hours on occupational activity. High-dynamic activity was more frequently entertained (10 [3-23] MET-hours/week) than high-static activity (0.1 [0-2] MET-hours/week).

After presentation, median activity decreased to 38 [14-106] MET-hours/week (p=0.021 compared to presentation). This was driven by a drastic decrease in occupational activities to 11 [0-73] MET-hours/week (p=0.022 compared to presentation), whereas recreational activities remained unchanged 14 [5-31] MET-hours/week (p=0.435 compared to presentation). While there was no significant change observed in high-static activities between presentation and follow-up (p=0.324), a reduction was noted in high-dynamic activities, p<0.001 compared to presentation. Boxplots of physical activity before and after presentation are provided in Supplemental Figure 2.

### Major Cardiomyopathy-related Events

In total, the primary endpoint (i.e. MCE) occurred in 40 (21%) individuals. This included 20 (11%) MVA events, 13 (7%) HF events, and 15 (8%) SRT events. Combinations of events occurred in eight (4%) individuals (three subjects with both MVA and HF, two subjects with both HF and SRT, and three subjects with MVA and SRT). None of the study subjects presented with MCE and median time to first MCE was 6.6 [2.2-11.9] years after presentation. There were no statistically significant differences in age (p=0.816) and sex (p=0.351) between those with and without MCE. In contrast, LV hypertrophy (21 vs. 14 mm, p<0.001) and LV ejection fraction (55 vs. 60%, p=0.011) were significantly different in those with vs. without MCE, respectively.

Figure 1A shows the effect of overall physical activity on the primary endpoint. As can be appreciated, no difference was observed for MCE occurrence between quartiles of overall physical activity measured in MET-hours/week (p=0.374).

**Figure 1.**
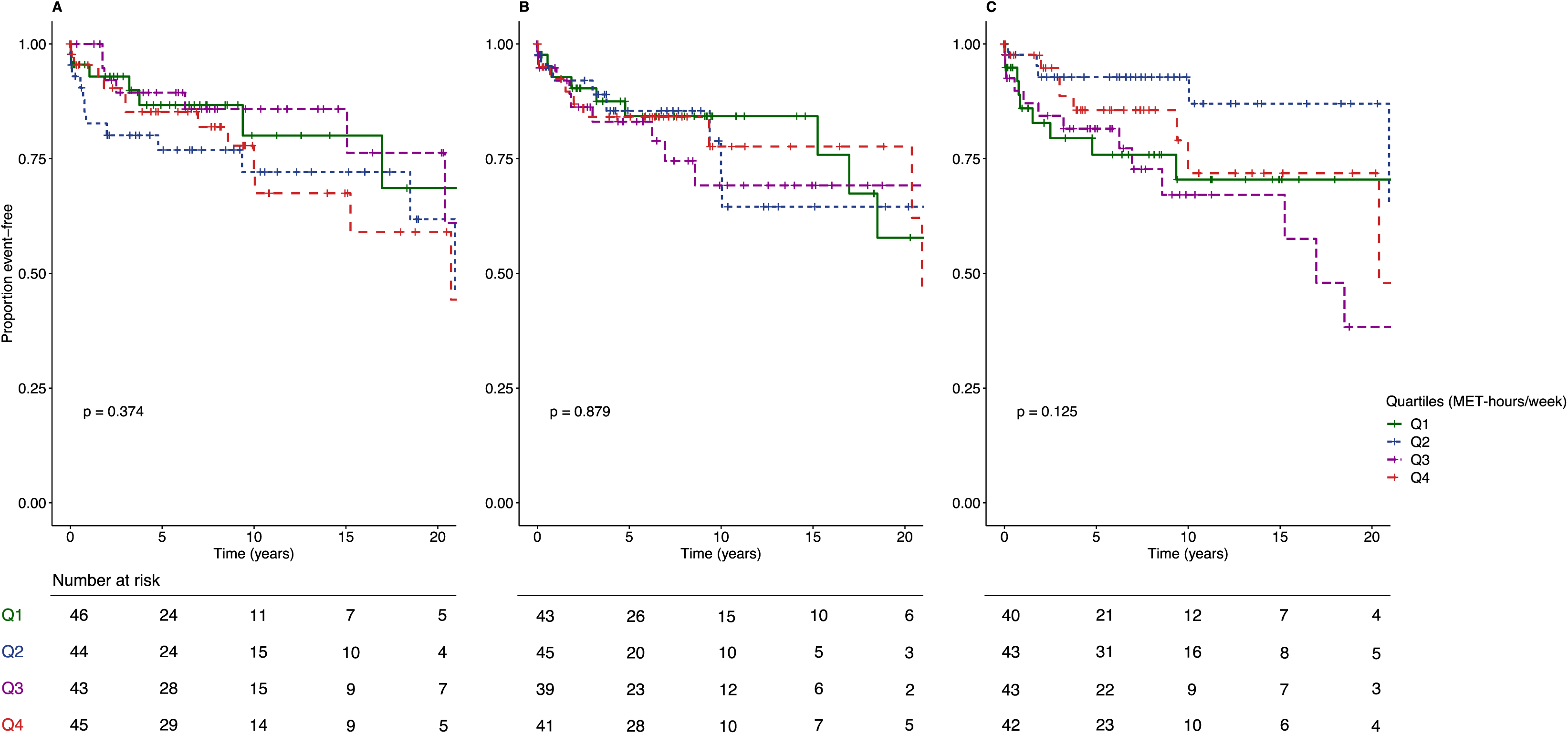
Survival free from major cardiomyopathy-related events. Kaplan-Meier curves showing cumulative survival free from MCE, stratified by quartiles of MET-hours/week, for **(A)** overall physical activity; **(B)** High-dynamic exercise, **(C)** High-static exercise. Abbreviations as in text.

Figure 1B and C show the effect of high-dynamic activity and high-static activity on MCE occurrence, respectively. As shown, there were no significant differences between quartiles of either activity category for MCE (p=0.879 for high-dynamic activity; and p=0.125 for high-static activity).

### Secondary Endpoints

#### Malignant ventricular arrhythmias

None of the individuals had experienced MVA at presentation. A total of 20 (11%) subjects experienced MVA by last follow-up, on average 6.8 [3.3-12.6] years after presentation. No differences were observed in age (p=0.998) or sex (p=0.063) between those with and without MVA.

Supplemental Figure 3A show the effect of overall physical activity and 3C the effect of high-static activity on MVA occurrence, respectively. As shown, there were no significant differences between quartiles of either activity category for MVA (p=0.130 for overall physical activity; and p=0.325 for high-static activity). As can be appreciated in Supplemental Figure 3B, an increased risk of MVA was observed among those engaged in the highest quartile of high-dynamic activities (p=0.041 compared to the lowest quartile of high-dynamic activities).

Assessing the impact of high-dynamic activity on the risk of MVA, a violation of linearity assumption of Cox proportional hazard was observed hence values of high-dynamic activity for this endpoint were dichotomized by its 75th percentile. As shown in Figure 2, high-dynamic exercise was associated with MVA when dichotomized by its 75th percentile (23 MET-hours/week, aHR 2.70, 95% CI 1.01-7.24, p=0.049).

**Figure 2.**
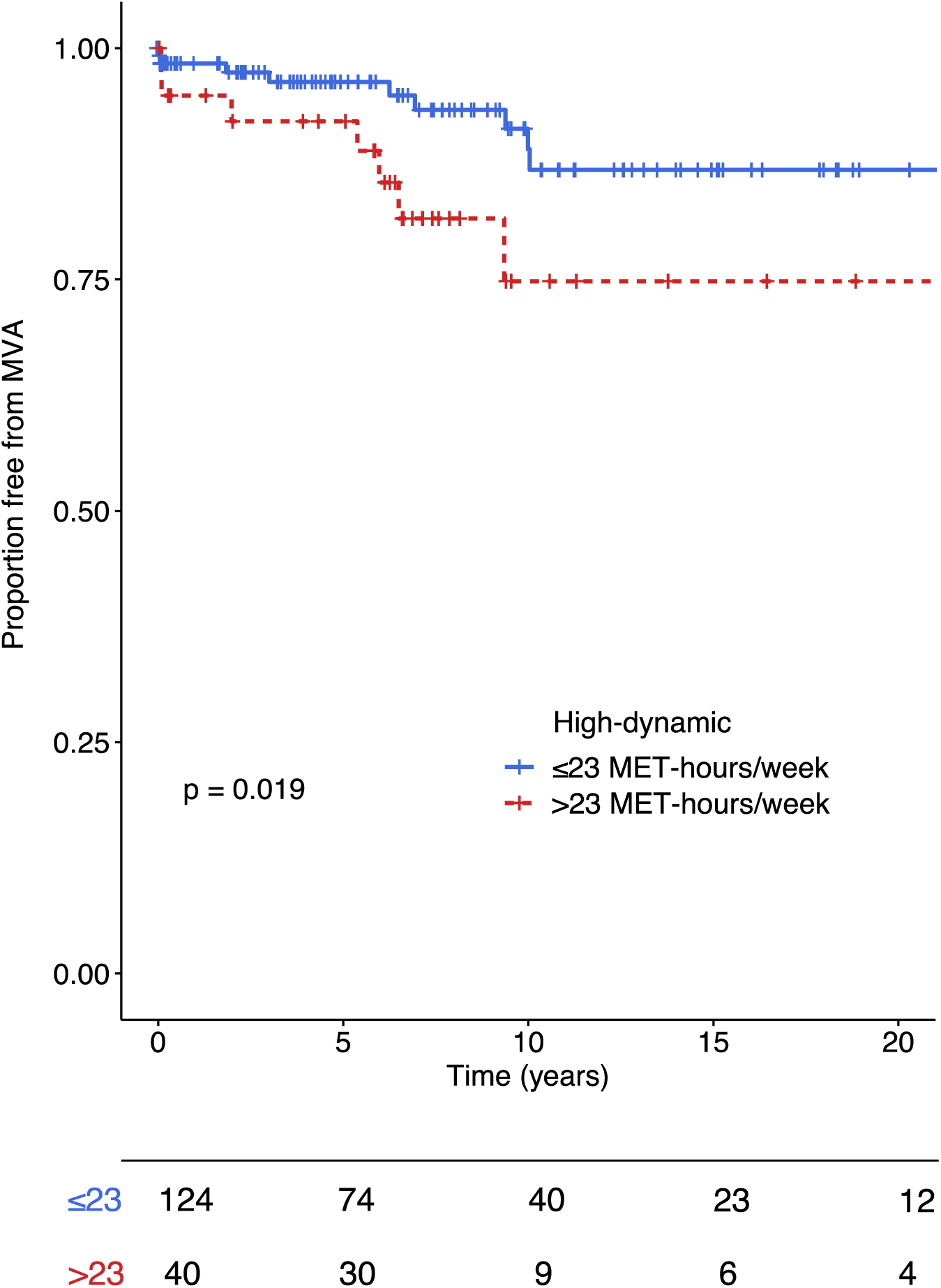
Survival free from malignant ventricular arrhythmia. Kaplan-Meier curves showing cumulative survival free from MVA, stratified by 75^th^ percentile MET-hours/week for high-dynamic exercise. Abbreviations as in text.

#### Heart failure events

None of the individuals had experienced HF events at presentation. A total of 13 (7%) subjects experienced HF events by last follow-up, on average 7.1 (3.2-13.9) years after presentation. No differences were observed in age (p=0.618) or sex (p=0.581) among those with and without HF events.

Supplemental Figure 4A shows the effect of overall physical activity on HF events. As can be appreciated, no difference was observed for HF occurrence between quartiles of overall physical activity measured in MET-hours/week (p=0.954).

Supplemental Figure 4B and C show the effect of high-dynamic activity and high-static activity on HF occurrence, respectively. There were no significant differences between quartiles of either activity category for HF events (p=0.490 for high-dynamic activity; and p=0.834 for high-static activity).

#### Septal reduction therapy

None of the individuals had experienced SRT at presentation. A total of 15 (8%) subjects experienced SRT by last follow-up, with a median 7.1 [3.3-13.9] years after presentation. No differences were observed in age (p=0.525) or sex (p=0.964) among those with and without SRT.

Supplemental Figure 5A shows the effect of overall physical activity on SRT. No difference was observed for SRT between quartiles of overall physical activity measured in MET-hours/week (p=0.539).

As shown in Supplemental Figure 5B (high-dynamic activity) and C (high-static activity), there were no significant differences between quartiles of either activity category for SRT (p=0.473 for high-dynamic activity; and p=0.072 for high-static activity).

#### Cardiomyopathy penetrance

A total of 104 (55%) subjects were diagnosed with cardiomyopathy (98 HCM, 3 DCM [possibly HCM in the “burned-out phase”], 2 CMP-NOS and 1 NCCM) at first clinical evaluation. During a median 7.4 [3.9-14.6] years of follow-up, 35 (19%) additional subjects developed cardiomyopathy, all of whom with an HCM diagnosis. This culminated to a total of 139 (74%) patients with cardiomyopathy by last follow-up.

Figure 3A shows the effect of overall physical activity on cardiomyopathy penetrance.

**Figure 3.**
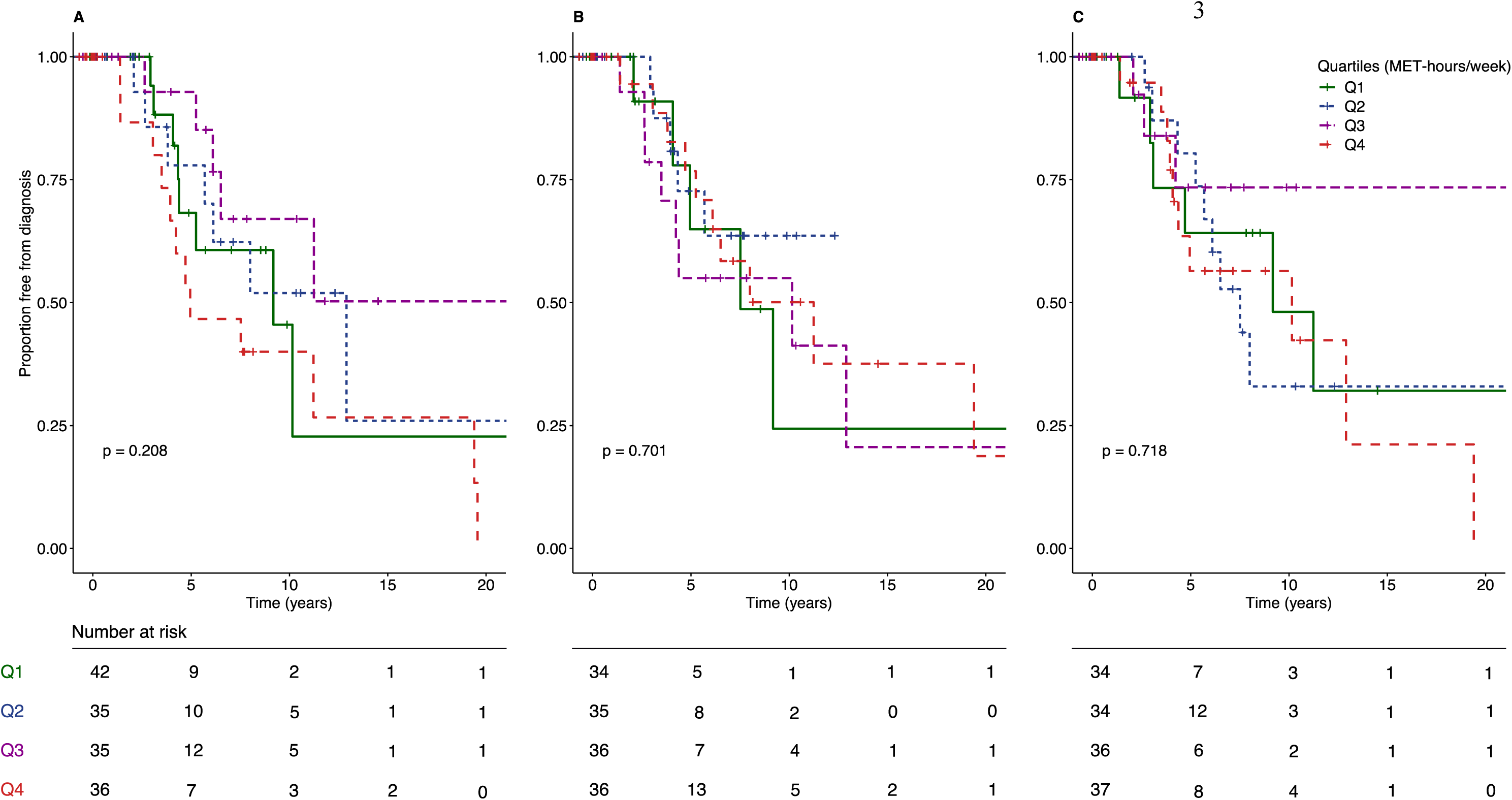
Cardiomyopathy penetrance. Kaplan-Meier curves showing cumulative survival free from cardiomyopathy penetrance, stratified by the quartiles of MET-hours/week, for **(A)** overall physical activity **(B)** high dynamic exercise and **(C)** high static exercise. Abbreviations as in text.

As can be appreciated, no difference was observed in cardiomyopathy penetrance between quartiles of overall physical activity measured in MET-hours/week (p=0.208).

Figure 3B and C show the effect of high-dynamic activity and high-static activity on cardiomyopathy penetrance, respectively. No significant differences between quartiles of either activity category for cardiomyopathy penetrance were identified (p=0.701 for high-dynamic activity; and p=0.718 for high-static activity).

### Sensitivity analysis

Our study population included a sizable number of subjects without phenotypic expression, which may influence our ability to pick up significant differences among groups. We therefore repeated the analyses solely in individuals with a cardiomyopathy diagnosis as sensitivity analysis. Clinical characteristics of subjects with a cardiomyopathy diagnosis are provided in Supplemental Table 2. In Supplemental Tables 3 and 4, it is shown that results were identical to the main results.

## Discussion

In this study, we evaluated the effects of physical activity stratified by type of activity on clinical outcomes and cardiomyopathy penetrance in 188 carriers of truncating *MYBPC3* founder variants. We assessed overall physical activity both in quartiles based on our data, and based on the American Heart Association’s recommended minimum for healthy adults (7.5 MET-hours/week). High-dynamic and high-static activities were classified as per Bethesda classification and evaluated in quartiles since there is no recommended cutoff for these types of activity.

Our findings revealed that neither overall physical activity nor high-static exercise increased the risk of MCE and cardiomyopathy penetrance. In contrast, when analyzing the combined endpoint by its separate components, we observed that high-dynamic exercise increased the risk of MVA when dichotomized by its 75^th^ percentile, which was equivalent to more than 23 MET-hours/week. For example, this is reached when an individual spends 50 minutes on vigorous swimming (9.8 METs/hour) for 3 days per week, or participates in 50 minutes of vigorous running (10 METs/hour) 3 days per week, or in 1.5 hours of vigorous soccer (10 METs/hour) 2 days per week.

### Physical activity and risk of MCE and cardiomyopathy penetrance

Physical activity has been traditionally considered as a risk factor for patients with HCM, which has resulted in conservative exercise recommendations. However, several recent studies have shown neutral or even beneficial effects of exercise in HCM. Saberi *et al*. assessed the impact of moderate-intensity exercise compared to usual activity in a preliminary randomized controlled trial including 136 patients with HCM. Not only did they not observe any MVA in either group, but they additionally observed a small but statistically significant increase in peak oxygen consumption in the moderate-intensity exercise group.^29^ Consistently, Klempfner et al. did not observe adverse events and showed increased functional capacity in a single-arm trial including 20 patients with symptomatic HCM who underwent a cardiac rehabilitation exercise program.^30^ Leveraging a nationwide population-based cohort, Kwon et al. observed decreases in all-cause and cardiovascular mortality for increasing tertiles of physical activity among 7666 patients with HCM during more than 5 years of follow-up.^16^ Likewise, in a retrospective, cross-sectional study, Andreassen et al. observed a positive association between exercise training during childhood and LV diastolic function in 121 patients with HCM and 66 carriers of HCM-associated genetic variants.^31^ Some authors have suggested that exercise restrictions have negative psychological effects in athletic patients with HCM, and may result in increased risks of cardiovascular disease resulting from a sedentary lifestyle.^32,33^ In sharp contrast to the abovementioned reassuring findings, however, in a small series Aengevaeren et al. found that HCM patients with increasing tertiles of physical activity were younger at the time of diagnosis and more frequently had a history of non-sustained ventricular tachycardia.^15^

A recent large prospective study shed light on this matter. In this study, Lampert et al. did not observe a higher rate of death or life-threatening arrhythmias among 1660 individuals with HCM exercising vigorously than those participating in moderate exercise.^34^ Of note, the authors used a cut-off of 360 MET-hours/year to define vigorous exercise, which is equivalent to approximately 7 MET-hours/week. This aligns closely with the AHA’s minimum recommended exercise level of 7.5 MET-hours/week for healthy adults. For example, this is reached when an individual spends one hour on vigorous bicycling (7.8 METs/hour) 1 day per week. This is a much lower cut-off than the 23 MET-hours/week that we used for the 75^th^ percentile of high-dynamic exercise, possibly explaining the observed difference between our studies. In line with the results of Lampert et al.,^34^ we found that overall physical activity and high-static exercise did not significantly increase the risk for any of our outcomes in carriers of truncating *MYBPC3* variants. Our study thus confirms and extends prior research, since our detailed data collection also allowed for sub-analyses stratified by specific types of exercise (i.e. by dynamic or static component), which may be more suitable to uncover possibly detrimental effects on clinical course in this patient population. Regardless of the available evidence, future prospective studies will be pivotal to better understand the interaction between exercise, HCM-related outcomes and general health benefits, in order to improve exercise recommendations for this generally young patient population.

### High-dynamic exercise and MVA

To the best of our knowledge, our study was the first to specifically assess high-dynamic and high-static exercise as potential predictors of cardiomyopathy-related outcomes. While we did not find an association between high-static exercise with any of our study outcomes, our data shows that those who participated in the highest quartile of high-dynamic exercise had increased risk of MVA suggesting that these activities should be entertained with caution. The exact mechanism underlying the association of high-dynamic exercise and MVA in these patients is not fully understood. Previous studies have proposed several factors such as heterogenous myocardial blood flow, myocardial disarray, associated abnormalities of the vascular response to exercise, electrolyte and sympathetic-vagal imbalance, and exercise-induced metabolic acidosis.^13,35–38^ More research in (pre-)clinical disease models may shed further light on this issue.

### High-static exercise and disease course

Our results showed that high-static exercise did not increase the risk of MCE and cardiomyopathy penetrance among *MYBPC3*-founder variant carriers. Whether high-static exercise can be considered safe, or whether there could be an effect of more intensive high-static exercise on disease course, cannot be determined from our data. Furthermore, caution is needed in those patients with LV outflow tract obstruction at rest or during exercise.^39–41^ Of note, out of 26 individuals with LV outflow tract obstruction in our study, one was engaged in frequent high-static exercise (>2 MET-hours/week), who experienced MCE. Therefore, we emphasize the importance of shared decision making for participation in more intensive static and also dynamic exercise. Future prospective studies are required to personalize exercise recommendations and to determine the optimal exercise threshold that is safe in this patient population.

### Limitations and Clinical Implications

To our knowledge, this is the first and largest interview-based exercise study in a genetically homogenous HCM population. Genotype-driven inclusion allowed us to assess cardiomyopathy penetrance as an outcome. Additionally, the design of the interview allowed us to assess specific categories of physical activity. This provides a foundation for future prospective studies to evaluate the effect of specific types of exercise on the disease course of patients with HCM and may contribute to finetuning exercise recommendations for these patients, balancing potential benefits of exercise with exercise-related risks.

Some limitations warrant consideration. The retrospective nature of this study results in ascertainment bias and missing data. Recall bias and social desirability bias may have resulted in over- or underreporting of physical activities. As carriers were required to be alive for the exercise interview, severely affected carriers that passed away before initiation of this study could not be included. Restricting inclusion to truncating *MYBPC3* variants may limit generalizability to the wider HCM population. Furthermore, the intensity of physical activity was only taken into account in the calculation of the number of MET-hours/week. However, different intensities of exercise could contribute to hypertrophic remodeling to differing degrees, e.g. vigorous exercise may be more detrimental than mild to moderate intensity exercise. Despite being the largest interview-based study in a genetically homogenous HCM population, sample size was still not sufficient to control for additional potential cofounding variables such as hypertension and body mass index. Additionally, we did not inquire about the use of anabolic steroids or other exercise-enhancing drugs and supplements, which could modify or confound the apparent effects of exercise in some carriers. Finally, in the absence of established cut-off values for high-static and high-dynamic exercise, we used data-driven cut-off values, which limit direct interpretation and may result in overfitting. Taken together, these limitations emphasize the importance of prospective validation of our results.

## Conclusions

In truncating *MYBPC3* founder variant carriers, overall physical activity and high-static exercise are not associated with an increased risk of MCE and cardiomyopathy penetrance. However, those who participated in the highest quartile of high-dynamic exercise had increased risk of MVA, suggesting that these activities should be entertained with caution. Taken together, these findings may contribute to improving exercise recommendations in HCM patients. We envision a more individualized approach and support the restrictions of very frequent high-intensity exercise in HCM patients. Future studies are needed to prospectively validate these findings.

## Clinical perspectives

### Competency in medical knowledge

The impact of different types of exercise on the risk of major cardiomyopathy-related events and cardiomyopathy penetrance in truncating *MYBPC3* founder variant carriers is unknown. We found that only those who participated in the highest quartile of high-dynamic exercise had increased risk of malignant ventricular arrhythmias; all other types of exercise did not result in increased risk of major cardiomyopathy-related events or cardiomyopathy penetrance. This suggests that while most exercise may be considered safe, very frequent high-dynamic exercise should be entertained with caution.

### Translational outlook

Further prospective research is necessary to validate these findings and determine safe thresholds and/or intensities of high-dynamic and high-static exercise for patients with cardiomyopathy, specifically HCM and carriers of pathogenic genetic variants.

## Data Availability

The data that support the findings of this study are available from the corresponding author upon reasonable request.

## Abbreviations and Acronyms

aHR: adjusted hazard ratio
HCM: hypertrophic cardiomyopathy
HF: heart failure
ICD: implantable cardioverter-defibrillator
LV: left ventricular
MCE: major cardiomyopathy-related events
MET: metabolic equivalent of task
MVA: malignant ventricular arrhythmia
SRT: septal reduction therapy
*MYBPC3*: myosin binding protein C3 gene

## Acknowledgements

We thank all study participants who made this study possible.

