## Supplemental Tables S1-S4; Supplemental Figures S1-S5 for "Recreational and occupational physical activity and risk of adverse events in truncating *MYBPC3* founder variant carriers"

*F. Hassanzada et al. (2024)*

*Supplemental Material*

Content

Supplemental Table 1. Subject characteristics of study population. 3

Supplemental Table 2. Characteristics of patients with a cardiomyopathy at first presentation stratified by exercise participation prior to presentation. 4

Supplemental Table 3. Sensitivity analyses. Risk of sports on primary endpoint (major cardiomyopathy-related events) 5

Supplemental Table 4. Sensitivity analysis. Risk of sports on secondary endpoints (MVA, HF, SRT) 6

Supplemental Figure 1. Classification of Sport 7

Supplemental Figure 2. Physical activity in study participants before and after presentation 8

Supplemental Figure 3. Malignant ventricular arrhythmia. 9

Supplemental Figure 4. Heart failure events. 10

Supplemental Figure 5. Septal reduction therapy. 11

**Supplemental Table 1.** Subject characteristics of study population.

| | **G+P- (n=84)** | **G+P+ (n=104)** | **P-value** |
|---|---|---|---|
| Age at presentation (years) | 40.7±14.8 | 45.5±14.5 | 0.027 |
| Male | 35 (41.7) | 73 (70.2) | <0.001 |
| Body surface area ($m^2$) | 1.9 (1.8-2.0) | 2.0 (1.8-2.2) | 0.013 |
| Proband | 12 (14.3) | 68 (65.4) | <0.001 |
| LV end-diastolic diameter (mm) | 47±5 | 45±8 | 0.380 |
| Left atrial diameter (mm) | 37±6 | 43±7 | <0.001 |
| Maximum wall thickness (mm) | 11±2 | 20±5 | <0.001 |
| LV ejection fraction | 60±5 | 58±12 | 0.104 |
| Phenotype | | | <0.001 |
| HCM | 0 (0.0) | 98 (94.2) | |
| DCM | 0 (0.0) | 3 (2.9) | |
| NCCM | 0 (0.0) | 1 (1.0) | |
| CMP NOS | 0 (0.0) | 2 (1.9) | |
| MET-hours/week before presentation spent on | | | |
| Overall physical activity | 49 (22-89) | 61 (31-121) | 0.126 |
| Exercise | 15 (6-36) | 15 (6-29) | 0.604 |
| Occupation | 24 (4-68) | 45 (6-103) | 0.097 |
| MET-hours/week after presentation spent on | | | |
| Overall physical activity | 39 (14-113) | 37 (15-100) | 0.799 |
| Exercise | 16 (7-32) | 11 (3-30) | 0.096 |
| Occupation | 11 (0-74) | 10 (0-68) | 0.749 |

Subject characteristics of study population stratified by absence or presence of a cardiomyopathy phenotype within one year after clinical presentation . Data are presented as mean ± standard deviation, median (interquartile range) or counts (percentage). Abbreviations: G+P-, genotype-positive phenotype-negative; G+P+, genotype-positive phenotype-positive. LV, left ventricular; HCM, hypertrophic cardiomyopathy; DCM, dilated cardiomyopathy; NCCM, non-compaction cardiomyopathy; CMP NOS, cardiomyopathy not otherwise specified; MCE, major cardiomyopathy-related events; MVA, malignant ventricular arrhythmia; HF, heart failure; SRT, septal reduction therapy; ICD implantable cardioverter-defibrillator; MET, metabolic equivalent task.

**Supplemental Table 2.** Characteristics of patients with a cardiomyopathy at first presentation stratified by exercise participation prior to presentation.

| | **Overall (n=98)** | **Quartile 1 (n=26)** | **Quartile 2 (n=26)** | **Quartile 3 (n=26)** | **Quartile 4 (n=26)** | **P-value** |
|---|---|---|---|---|---|---|
| Male sex | 73 (70.2) | 16 (61.5) | 16 (61.5) | 19 (73.1) | 22 (84.6) | 0.208 |
| Body surface area ($m^2$) | 2.0 (1.9-2.2) | 1.9 (1.9-2.2) | 2.2 (2.1-2.3) | 2.0 (1.7-2.0) | 2.0 (1.9-2.2) | 0.354 |
| Age at presentation (years) | 45.5 (14.5) | 45.9 (11.7) | 43.6 (18.9) | 45.9 (12.6) | 46.6±4 | 0.886 |
| LV end-diastolic diameter (mm) | 45±8 | 43±6 | 46±8 | 44±7 | 49±9 | 0.288 |
| Left atrial diameter (mm) | 43±7 | 42±4 | 42±7 | 48±9 | 40±6 | 0.101 |
| Maximum wall thickness (mm) | 20±5 | 21±4 | 21±6 | 20±5 | 19±5 | 0.397 |
| LV ejection fraction (%) | 58±12 | 59±7 | 61±14 | 58±7 | 54±17 | 0.350 |

Subject characteristics of the 104 carriers with a cardiomyopathy at first presentation, stratified by average MET-hours/week of overall physical activity quartiles before first presentation. Study outcomes, ICD implantations and atrial arrhythmia occurring within the first year after first presentation were included in the above counts. Values are presented as mean ± standard deviation, median (interquartile range) or counts (percentage). Abbreviations as in Supplemental Table 1.

**Supplemental Table 3.** Sensitivity analyses. Risk of sports on primary endpoint (major cardiomyopathy-related events)

| | **HR*** | **95% CI** | **P-value** |
|---|---|---|---|
| Overall physical activity (per 7.5 MET-hours/week of AHA minimum recommended exercise level) | 1.00 | 1.00-1.00 | 0.694 |
| High dynamic (per 1 MET-hours/week) | 1.00 | 0.99-1.02 | 0.602 |
| High static (per 1 MET-hours/week) | 0.98 | 0.92-1.03 | 0.398 |

*Corrected for sex. Abbreviation as in text.

**Supplemental Table 4.** Sensitivity analysis. Risk of sports on secondary endpoints (MVA, HF, SRT)

| | **HR*** | **95% CI** | **P-value** |
|---|---|---|---|
| **MVA** | | | |
| Overall physical activity (per 7.5 MET-hours/week of AHA minimum recommended exercise level) | 1.00 | 1.00-1.00 | 0.489 |
| High dynamic (Dichotomized by median 10 MET-hours/week) | 3.18 | 1.07-9.47 | 0.038 |
| High dynamic (Dichotomized by 75th percentile 23 MET-hours/week) | 3.43 | 1.27-9.25 | 0.015 |
| High static (per 1 MET-hours/week) | 0.99 | 0.93-1.05 | 0.706 |
| **HF** | | | |
| Overall physical activity (per 7.5 MET-hours/week of AHA minimum recommended exercise level) | 1.00 | 1.00-1.00 | 0.946 |
| High dynamic (per 1 MET-hours/week) | 1.02 | 0.99-1.04 | 0.272 |
| High static (per 1 MET-hours/week) | 0.95 | 0.83-1.09 | 0.486 |
| **SRT** | | | |
| Overall physical activity (per 7.5 MET-hours/week of AHA minimum recommended exercise level) | 1.00 | 1.00-1.00 | 0.709 |
| High dynamic (per 1 MET-hours/week) | 0.98 | 0.94-1.02 | 0.352 |
| High static (per 1 MET-hours/week) | 0.99 | 0.92-1.08 | 0.870 |

*Corrected for sex. Abbreviation as in text

**Supplemental Figure 1.** Classification of Sport

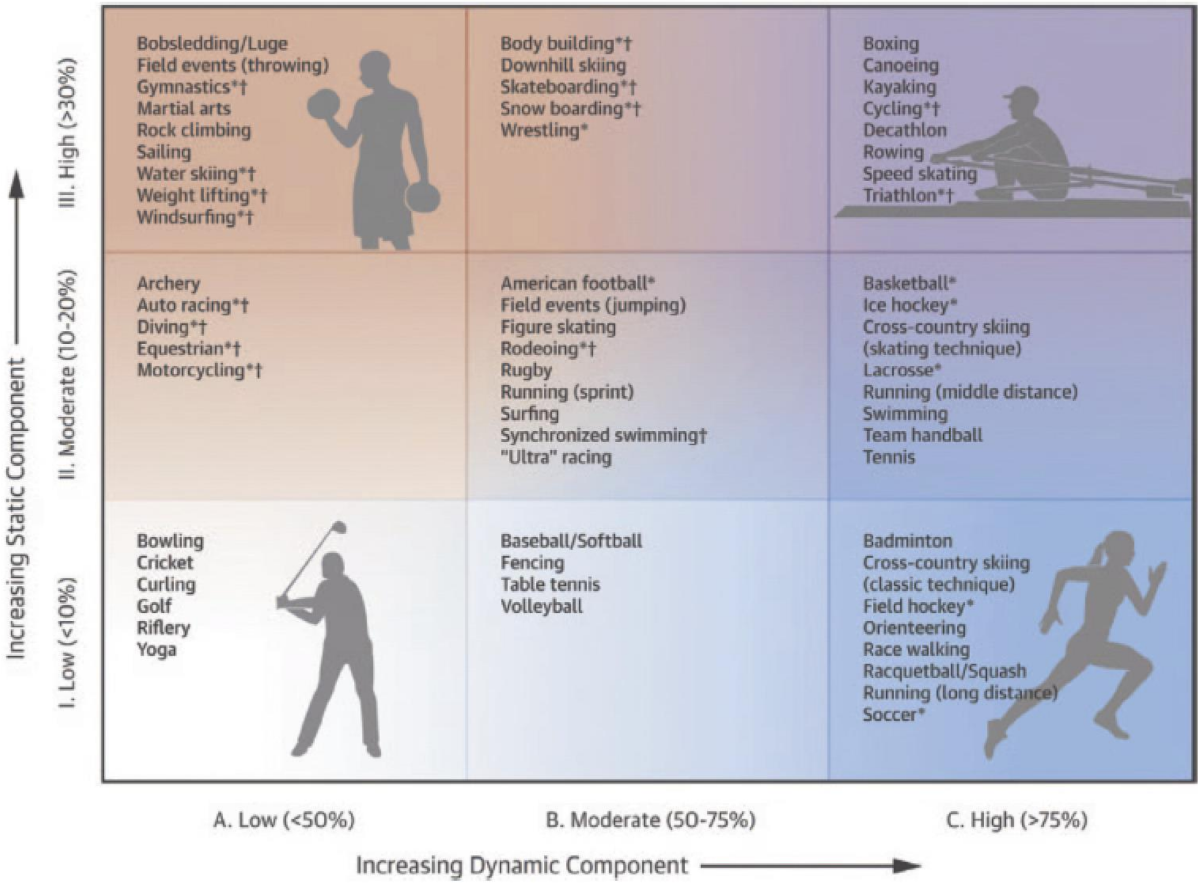


Figure from: Levine BD, Baggish AL, Kovacs RJ, Link MS, Maron MS, Mitchell JH. Eligibility and disqualification recommendations for competitive athletes with cardiovascular abnormalities: task force 1: classification of sports: dynamic, static, and impact a scientific statement from the American Heart Association and American College of Cardiology. J Am Coll Cardiol 2015; 66:2350–5. Reproduced with permission.

**Supplemental Figure 2.** Physical activity in study participants before and after presentation.

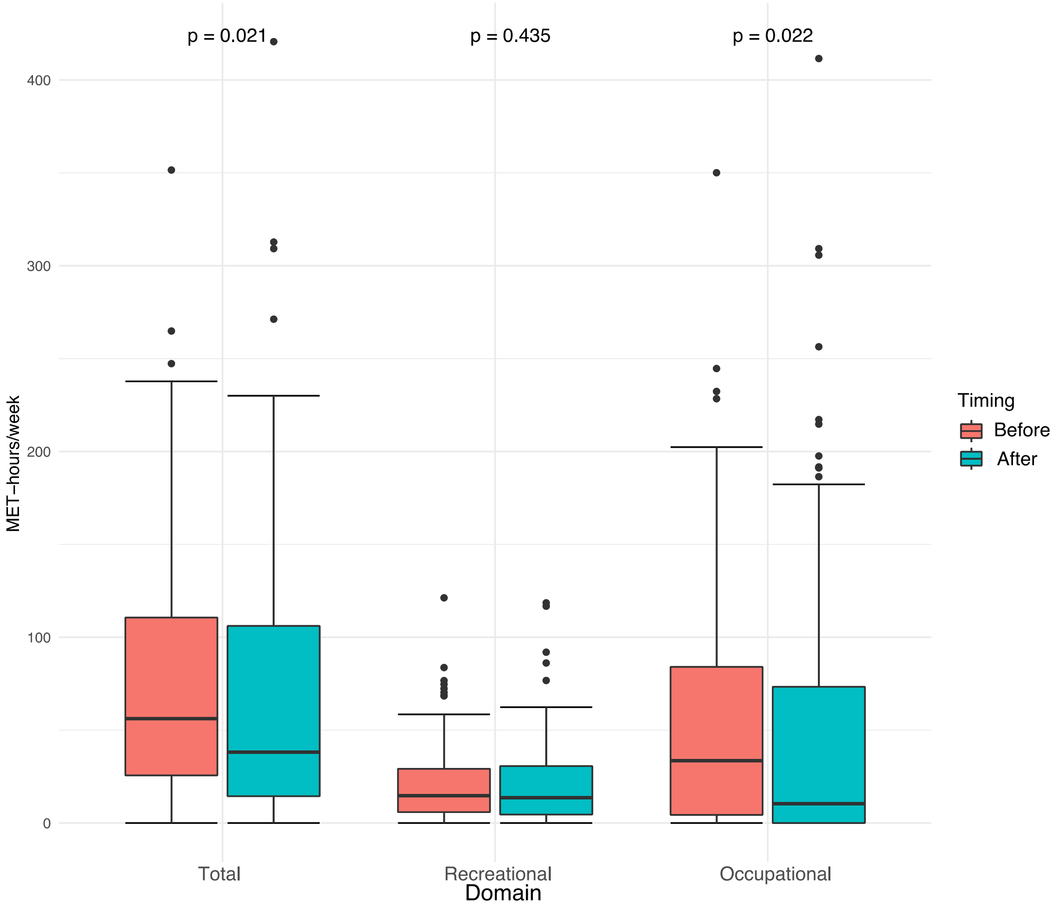


Boxplots depicting MET-hours/week of physical activity of participants before presentation in red and after presentation in blue. Physical activity is further specified into exercise and occupational activity. P-values were obtained using Mann-Whitney U test.

**Supplemental Figure 3.** Malignant ventricular arrhythmia.

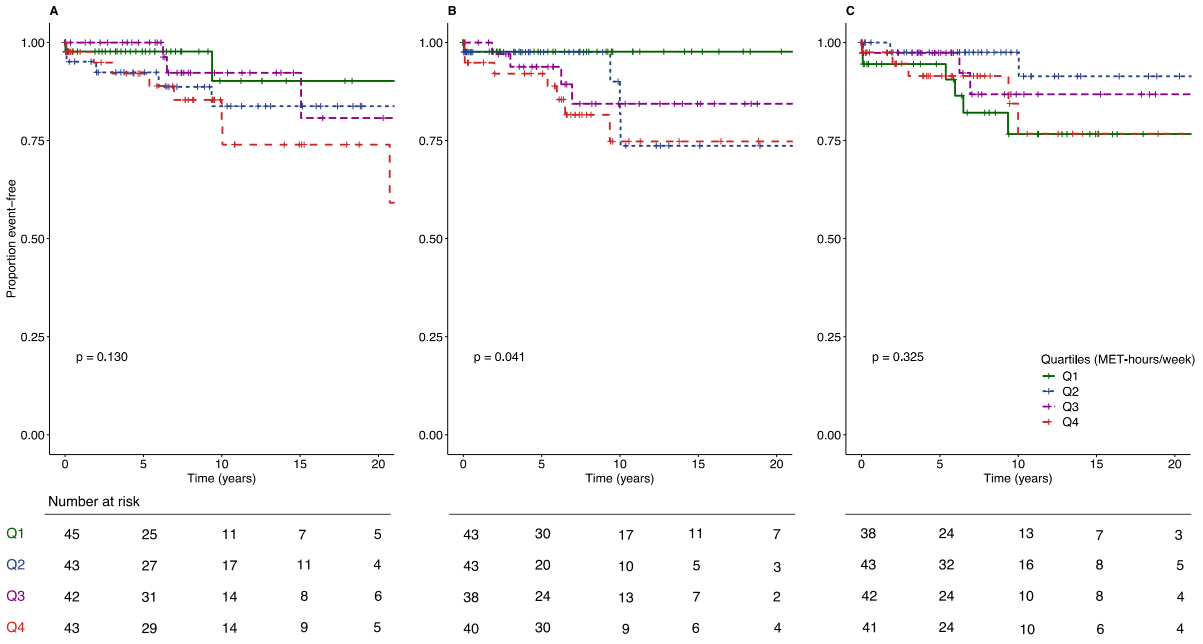


Kaplan-Meier curves showing cumulative survival free from MVA events, stratified by the quartiles MET-hours/week, for **(A)** overall physical activity **(B)** high dynamic exercise and **(C)** high static exercise. Abbreviations as in text.

**Supplemental Figure 4.** Heart failure events.

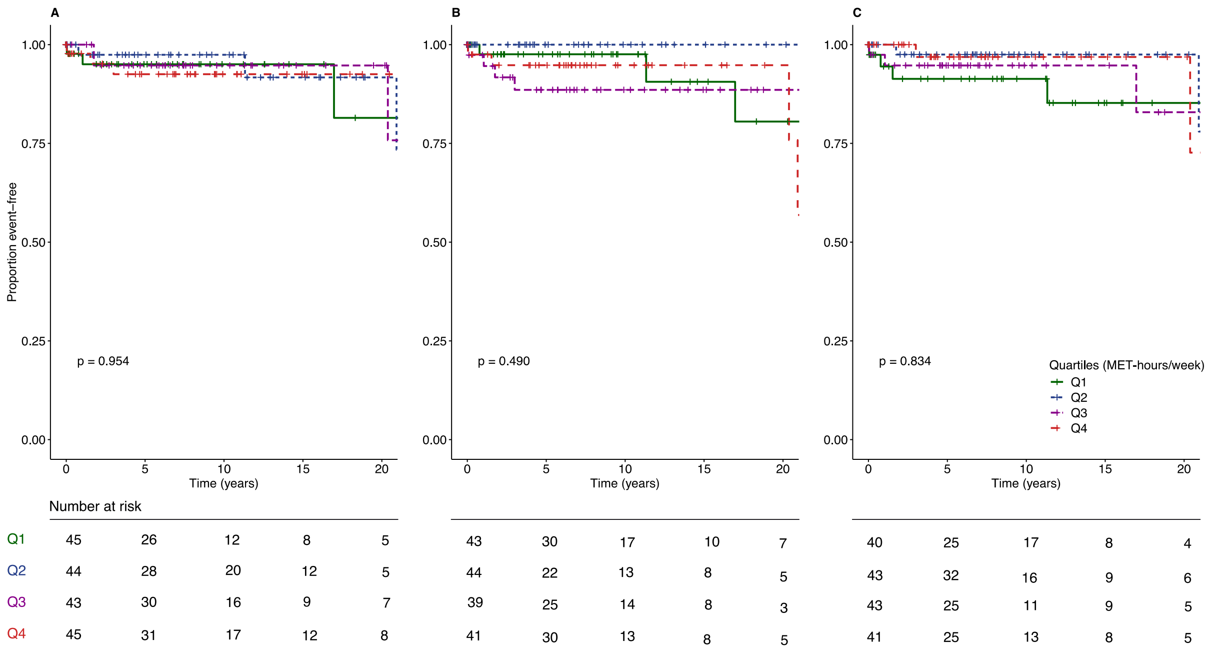


Kaplan-Meier curves showing cumulative survival free from HF events, stratified by the quartiles MET-hours/week, for **(A)** overall physical activity **(B)** high dynamic exercise and **(C)** high static exercise.

**Supplemental Figure 5.** Septal reduction therapy.

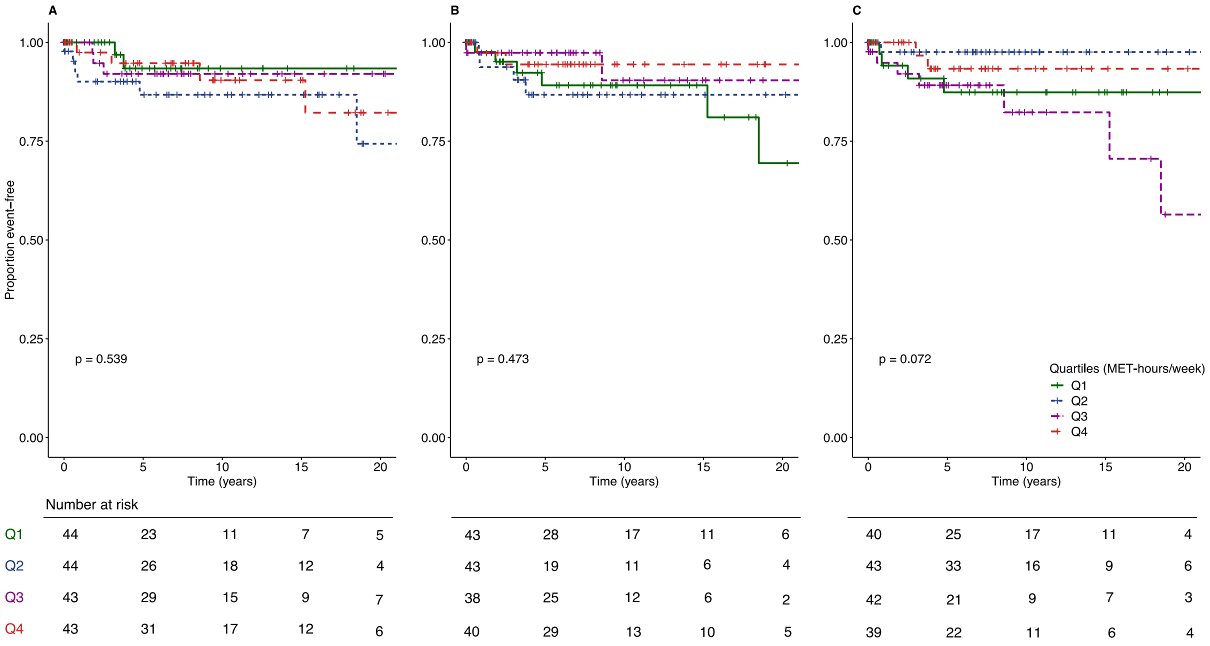


Kaplan-Meier curves showing cumulative survival free from septal reduction therapy, stratified by the quartiles MET-hours/week, for **(A)** overall physical activity **(B)** high dynamic exercise and **(C)** high static exercise.
